# Independent effect and joint effect of polygenic liabilities for schizophrenia and bipolar disorder on cognitive aging and education attainment

**DOI:** 10.1101/2022.03.14.22272327

**Authors:** Chi-Shin Wu, Chia-Lin Hsu, Mei-Chen Lin, Mei-Hsin Su, Yen-Feng Lin, Chia-Yen Chen, Po-Chang Hsiao, Yi-Jiun Pan, Pei-Chun Chen, Yen-Tsung Huang, Shi-Heng Wang

## Abstract

To elucidate the specific and shared genetic background of schizophrenia (SCZ) and bipolar disorder (BPD) to better understand their nosology, this study explored the independent and joint effects of polygenic liabilities for SCZ and BPD on cognitive aging and educational attainment among a collection of 80318 unrelated community participants from the Taiwan Biobank. Using the Psychiatric Genomics Consortium meta-analysis as a discovery sample, we calculated the polygenic risk score (PRS) for SCZ (PRS_SCZ_) and BPD (PRS_BPD_), shared PRS between SCZ and BPD (PRS_SCZ+BPD_), and SCZ-specific, differentiated from BPD, PRS (PRS_SCZvsBPD_). Based on the sign concordance of the susceptibility variants with SCZ and BPD, PRS_SCZ_ was split into PRS_SCZ_concordant_ and PRS_SCZ_discordant_ and PRS_BPD_ was split into PRS_BPD_concordant_ and PRS_BPD_discordant_. Linear regression models were used to estimate the association with cognitive aging as measured by the Mini-Mental State Examination (MMSE) in individuals aged ≥ 60 years. Ordinal logistic regression models were used to estimate the association with educational attainment. PRS_SCZ_ was negatively associated with MMSE (beta=-0.063, p<0.001), while PRS_BPD_ was positively associated with MMSE (beta=0.04, p=0.01). A larger difference between PRS_SCZ_ and PRS_BPD_ was associated with lower MMSE scores (beta=-0.052, p<0.001). Both PRS_SCZ_concordant_ and PRS_SCZ_discordant_ were negatively associated with MMSE scores, without a synergistic effect. There was a complex interaction between PRS_BPD_concordant_ and PRS_BPD_discordant_ on the MMSE scores. PRS_SCZ+BPD_ (beta=-0.09, p=0.01) and PRS_SCZvsBPD_ (beta=-0.13, p<0.001) predicted a decrease in MMSE scores during the follow-up. PRS_SCZ,_ PRS_BPD_, and PRS_SCZ+BPD_ were positively associated with educational attainment, whereas PRS_SCZvs BPD_ was negatively associated with educational attainment. This study provides evidence for the contrasting effect of polygenic liabilities for SCZ and BPD on cognitive aging and partially supports the hypothesis that the heterogeneity of SCZ and the positive association of polygenic liability for SCZ with education might be attributed to the shared part with BPD.

## Introduction

Major psychiatric disorders are differentiated on the basis of symptom patterns and course of illness from patient descriptions and observations of behavior made by clinicians or reported by informants. Clinical features, such as psychosis and cognitive impairment, have also been known to transcend diagnostics. The boundaries between psychiatric disorders are not clear-cut.^1, 2^ A patient could have heterogeneous features, and the boundaries between psychiatric disorders can be blurred. Whether schizophrenia (SCZ) and bipolar disorder (BPD) are clinical features of discrete or shared causative processes is unclear.

The genetic architecture is crucial to the causation of psychiatric disorders,^3^ and genetic studies have been used to assess potential overlaps.^4, 5^ Over 100 genome-wide significant loci shared between SCZ and BPD have been identified.^6^ Using a polygenic risk score approach to explore genetic overlap,^7^ a cross-disorder association between SCZ and BPD was revealed.^4^ Beyond shared polygenic risk, a recent large genome-wide association study (GWAS) further explored the shared symptoms between SCZ and BPD driven by the same underlying polygenic profiling^6^ and found polygenic profiles that significantly correlate with symptoms of both disorders. However, GWAS has also identified specific loci for distinguishing between BPD and SCZ.^6^

Cognitive deficits are the core symptoms of schizophrenia,^8-11^ usually occur many years before the onset of schizophrenia,^8, 12-14^ and may worsen after psychosis is present.^8^ The evidence for premorbid cognitive deficits in patients with BPD has been equivocal. After illness onset, cognitive function declines in patients with SCZ or BPD, but the magnitude of decline in cognition is greater in patients with SCZ than in patients with BPD.^15-17^. Up to 33.5 % of the heritable liability for global SCZ might be mediated through cognitive functions.^18^ Several genetic loci related to SCZ are also associated with poor cognitive performance.^19^ A Mendelian randomization analysis showed that there is a bidirectional causal association between SCZ and intelligence. In contrast, there is no significant causal association between BPD and intelligence.^20^ A negative correlation between general cognitive function and genetic factors differentiating SCZ from BPD has been revealed using GWAS summary data.^21^ It has been proposed that the diagnosis of SCZ gathers at least two disease subtypes.^22^ One subtype has been suggested to be similar to BPD and have high intelligence, and the other who are independent of BPD manifested cognitive impairment. A higher polygenic risk for SCZ is associated with lower cognitive performance in the general population^23-27^ and clinical samples.^27-29^ However, the genetic evidence for the association between SCZ/BPD and cognitive aging among elderly individuals is limited. For example, it is unclear whether polygenic liabilities for SCZ and BPD as well as their joint effect are associated with cognitive aging.

In patients with SCZ and BPD, higher educational attainment is associated with a lower degree of intelligence decline from the premorbid phase to after illness onset.^17^ However, illness onset may interrupt the educational course. Education attainment is lower in patients with SCZ or BPD.^30-32^ Poor primary school performance is also associated with subsequent risk of SCZ.^33^ Contrarily, genetic studies using GWAS summary data showed that there is a positive genetic correlation between education attainment and the two disorders, and the coefficient of correlation is stronger for BPD than for SCZ.^21, 34^ Both higher polygenic liabilities for SCZ and BPD are also associated with higher education attainment.^25, 35^ However, an association between higher polygenic liability for SCZ and lower education attainment has been reported in UK Biobank.^36^ Besides, SCZ-specific genetic background differentiated from BPD has a negative genetic correlation with education attainment.^21^ A Danish population-based study showed that the observed association between primary school performance and subsequent risk of SCZ cannot be explained by polygenic liability for SCZ.^33^ The evidence for a genetic association between SCZ, BPD, and education attainment remains conflicting, and studying the joint effect for polygenic liabilities for SCZ and BPD would provide novel implications.

To elucidate the specific and shared genetic background of SCZ and BPD to better understand the nosology of psychiatric disorders, this study aimed to explore the independent, shared, and contrasting effects of polygenic liabilities for SCZ and BPD on cognitive aging and education attainment among a large collection of community samples from the Taiwan Biobank.

## Material and methods

### Study samples and measurements

The study participants were recruited from the Taiwan Biobank,^37, 38^ the largest government-supported biobank in Taiwan since 2012, with genomic data and repeated measurements of a wide range of phenotypes collected, with an expected final sample size of 200 000. The Taiwan Biobank recruits community-based participants aged 30–70 years, without a history of cancer. The recruitment and sample collection procedures were approved by the internal review board of the Taiwan Biobank. Each participant signed an approved informed consent form, provided blood samples, and participated in face-to-face interviews. This study was approved by the Central Regional Research Ethics Committee of the China Medical University, Taichung, Taiwan (CRREC-108-30).

The questionnaire was conducted through face-to-face interviews with each participant. The questionnaire included demographic information, socio-economic status, self-reported disease status, and Mini-Mental State Examination (MMSE). Educational attainment was classified as illiteracy, self-study, elementary school, junior high school, senior high/vocational school, university/college, and master and above. The MMSE, the most commonly used tool for testing cognitive aging, was measured in subjects aged > 60 years. Cognitive deficit was defined as an MMSE score of < 24. Some individuals were followed up; hence, the MMSE change during follow-up could be calculated.

### Genetic analysis and quality control

This study included 95 238 individuals for whom genome-wide genotyping was performed. Genome-wide genotyping was performed using the custom Taiwan Biobank chips and run on the Axiom Genome-Wide Array Plate System (Affymetrix, Santa Clara, CA, USA); 26 274 participants were genotyped on the TWBv1 chip and 68 964 participants were genotyped on the TWBv2 chip. Quality control of the two batches was conducted separately before imputation. Initial quality control included the exclusion of individuals with more than 5 % missing variants and kinship pairs, excluding variants with a call rate < 5 %, minor allele frequency < 0.001, and deviation from Hardy–Weinberg equilibrium with P < 1 × 10^−6^. To exclude cryptic relatedness, we estimated identity by descent (IBD) sharing coefficients, PI-HAT = probability (IBD = 2) + 0.5 × probability (IBD = 1), between any two participants and excluded one individual from a pair with PI-HAT > 0.1875. A total of 80 318 unrelated individuals remained in the study. We analyzed the population stratification using principal components derived from genome-wide genotypic data. No population structure or heterozygosity outliers were found. We used the 504 EAS panel from 1000 Genomes Project^39^ and the 973 TWB panel from whole-genome sequencing in TWB participants as the reference panel to impute genotype with IMPUTE2 for the two chips separately (16 537 709 variants for TWBv1 and 16 222 535 variants for TWBv2), and then retained variants with imputation info score > 0.7 (13 803 712 variants for TWBv1 and 13 586 691 variants for TWBv1). A total of 12 605 051 variants were available in both chips and kept for subsequent PRS calculations. The chip version was adjusted for PRS-association analyses.

### Polygenic risk score calculation

Data from the Psychiatric Genomics Consortium (PGC) meta-analysis were used as discovery samples to identify susceptibility variants for SCZ and BPD. The shared polygenic risk score (PRS) between SCZ and BPD (PRS_SCZ+BPD_) was derived based on a GWAS consisting of 53 555 SCZ + BPD patients and 54 065 controls.^6^ The SCZ-specific PRS differentiated from BPD (PRS_SCZvsBPD_) was derived based on a GWAS consisting of 23 585 patients with SCZ and 15 270 patients with BPD.^6^

PRS for SCZ (PRS_SCZ_) was derived from GWAS for SCZ with 67 390 cases and 94 015 controls.^40^ The PRS for BPD (PRS_BPD_) was derived from GWAS for BPD with 41 917 cases and 371 549 controls.^41^

To consider at least two subtypes of SCZ, one resembling BPD and the other independent of BPD, we split PRS_SCZ_ into two scores based on sign-concordant^22^ of the variants with SCZ and BPD, to capture the genetic heterogeneity of SCZ. These variants that had concordant signs for SCZ and BPD (risk and risk or protective and protective on both traits) were combined into a PRS for SCZ concordant (PRS_SCZ_concordant_), and the remaining variants that had discordant signs (risk, protective, protective, and risk for both traits) were combined into a PRS for SCZ discordant (PRS_SCZ_discordant_). In addition, we split PRS for BPD into PRS for BPD concordant (PRS_BPD_concordant_) and PRS for BPD discordant (PRS_BPD_discordant_).

To remove variants in linkage disequilibrium, variants were clumped with a pairwise R^2^ threshold of 0.5 and a sliding window size of 250 kb. Sets of variants with p-values below different thresholds for the association test with SCZ were defined as 1, 0.5, 0.1, and 0.05, which were the thresholds that reached a good balance between true-associated and null signals to capture heritability. A threshold of 1 for PRS was used in the main analyses, and thresholds of 0.5, 0.1, and 0.05 were used for sensitivity analyses. For each subject in the Taiwan Biobank, PRS was calculated using PLINK and normalized to a Z-score. The variance explained by the association of PRSs, including PRS_SCZ_, PRS_BPD_, PRS_SCZ+BPD_, PRS_SCZvsBPD_, PRS_SCZ_concordant_, PRS_SCZ_discordant_, PRS_BPD_concordant_, and PRS_BPD_discordant_, with the corresponding diseases, including SCZ and BPD, was examined.

### Statistical analysis

The distribution of demographic factors, educational attainment, and MMSE was described by number and percentage, or mean and standard deviation (SD), according to the characteristics of the data. To explore the association between PRS and MMSE scores at baseline and MMSE changes during follow-up, a linear regression model with adjustment for sex, age, birth cohort, education attainment, batch version, and top 20 population stratification dimensions was performed. To test the association with cognitive deficits (MMSE score < 24) at baseline, a logistic regression model with the same adjustments was performed. To explore the association with educational attainment, including five categories (from elementary school to master and above), an ordinal logistic regression model with adjustment for gender, age, birth cohort, batch version, and the top 20 population stratification dimensions was performed.

Several model settings were tested to explore the complexity of polygenic liabilities for SCZ and BPD. Model 1 was used to test the independent effects of PRS_SCZ_ and PRS_BPD_. Model 2 tested the combined effect of PRS_SCZ_ and PRS_BPD_ by examining the mean of PRS_SCZ_ and PRS_BPD_, and the difference (PRS_SCZ_ - PRS_BPD_). Model 3 tested the shared liabilities between SCZ and BPD (PRS_SCZ+BPD_). Model 4 tested the SCZ-specific risk contrast to BPD (PRS_SCZvsBPD_).

We further analyzed two PRS jointly on a bivariate continuous scale using thin-plate splines, which is a two-dimensional smoother with the estimated curvature in local regions moving over the two-dimensional (bivariate) surface of two PRS. We analyzed MMSE by PRS_SCZ_ and PRS_BPD_ jointly, PRS_SCZ_concordant_ and PRS_SCZ_discordant_ jointly, and PRS_BPD_concordant_ and PRS_BPD_discordant_ jointly.

The overall significance level was set at p < 0.05. All statistical analyses were performed using SAS 9.4.

## Results

Among 80 318 unrelated individuals, 141 reported having SCZ and 510 reported having BPD. The variance explained and the p-value of the association of the PRS at a threshold of 1 with SCZ or BPD are shown in Table 1. For predicting SCZ, PRS_SCZ_ and PRS_SCZ_concordant_ explained the most variance (3.74 % and 3.63 %, respectively), PRS_SCZ_discordant_ explained only 0.79 %. PRS_BPD_concordant_ explained more variance in SCZ than PRS_BPD_ (1.08 % vs. 0.89 %). For predicting BPD, PRS_BPD_concordant_ led to a larger explained variance than PRS_BPD_ (0.38 % vs. 0.28 %), while PRS_BPD_discordant_ explained little variance in BPD. The results for PRS thresholds of 0.5, 0.1, and 0.05 are shown in Supplementary Table 1; the pattern was similar.

**Table 1.** The association of polygenic risk score (PRS), at a threshold of 1, for schizophrenia and bipolar disorder with the corresponding diseases in 80318 unrelated participants form the Taiwan Biobank data.

| PRS | Schizophrenia |  |  | Bipolar disorder |  |  |
| --- | --- | --- | --- | --- | --- | --- |
|  | 141 cases & 80177 controls |  |  | 510 cases & 79808 controls |  |  |
|  | OR | p-value | R<sup>2a</sup> (%) | OR | p-value | R<sup>2a</sup> (%) |
| SCZ | 2.10 | <0.001 | 3.74% | 1.22 | <0.001 | 0.33% |
| BPD | 1.43 | <0.001 | 0.89% | 1.20 | <0.001 | 0.28% |
| SCZ & BPD shared | 1.52 | <0.001 | 1.18% | 1.20 | <0.001 | 0.29% |
| SCZ specific | 1.19 | 0.044 | 0.20% | 0.97 | 0.521 | 0.01% |
| SCZ concordant | 2.08 | <0.001 | 3.63% | 1.23 | <0.001 | 0.35% |
| SCZ discordant | 1.40 | <0.001 | 0.79% | 1.08 | 0.095 | 0.05% |
| BPD concordant | 1.74 | <0.001 | 2.08% | 1.24 | <0.001 | 0.38% |
| BPD discordant | 0.83 | 0.025 | 0.25% | 1.01 | 0.754 | 0.00% |

<sup>a</sup> Increase in Nagelkerke pseudo R<sup>2</sup> when adding the PRS into the model including gender, age, batch version, and 20 population stratification dimensions.

In the PRS association analyses for MMSE and educational attainment, individuals with self-reported SCZ or BPD (n = 639; 12 cases with both diseases) and those with missing values for education attainment (n = 28) were excluded. A total of 79 651 samples remained, and the distribution of demographics is shown in Table 2. There were 50 447 females (63.3 %), with a mean age of 50. The most common educational attainment was university (46.3 %). The proportion of illiterate (0.2 %) and self-study (0.1 %) participants was low. They were, therefore, not included in the analyses for the association with educational attainment. The MMSE was available for 20 276 individuals aged > 60 years. The mean MMSE score was 27.4, and 1496 participants (7.4 %) were categorized as having cognitive deficits. Follow-up data for MMSE were available for 4248 individuals, and the mean change in MMSE score was 0.3, which may be due to the learning effect.

**Table 2.** Demographic characteristics among 79,651 unrelated individuals without schizophrenia and bipolar disorder form the Taiwan Biobank data.

| Variable | n (%) |
| --- | --- |
| Gender |  |
| Female | 50447 (63.3) |
| Male | 29204 (36.7) |
| Age at baseline |  |
| 30-40 | 16746 (21.0) |
| 41-50 | 19694 (24.7) |
| 51-60 | 25197 (31.6) |
| 61-70 | 18014 (22.6) |
| Mean ± SD | 50.0 ± 10.7 |
| Birth cohort |  |
| <1950 | 5853 (7.4) |
| 1950-1959 | 23446 (29.4) |
| 1960-1969 | 22636 (28.4) |
| 1970-1979 | 18564 (23.3) |
| 1980-1989 | 9152 (11.5) |
| Education attainment |  |
| Illiterate | 123 (0.2) |
| Self-study | 48 (0.1) |
| Elementary school | 4128 (5.2) |
| Junior high school | 6191 (7.8) |
| Senior high/Vocational school | 24363 (30.6) |
| University/College | 36914 (46.3) |
| Master and above | 7884 (9.9) |
| MMSE at baseline (n=20,276) |  |
| <24 | 1496 (7.4) |
| ≥24 | 18780 (92.6) |
| Mean ± SD | 27.4 ± 2.5 |
| MMSE Change (n=4,248) |  |
| Mean ± SD | 0.3 ± 2.4 |

SD: standard deviation

The results of the association of polygenic liabilities for SCZ and BPD with MMSE and educational attainment under different models are shown in Table 3. In the model testing the independent effect of polygenic liabilities for SCZ and BPD, PRS_SCZ_ was negatively associated with MMSE (beta in per SD increase in PRS = -0.063, p < 0.001), while PRS_BPD_ was positively associated with MMSE (beta = 0.04, p = 0.01). The combined effect model suggested that the larger difference between PRS_SCZ_ and PRS_BPD_ was associated with lower MMSE (beta = -0.052, p < 0.001). Both shared risks between SCZ and BPD, PRS_SCZ+BPD_, and SCZ-specific PRS, PRS_SCZvsBPD_, were not associated with baseline MMSE.

**Table 3.** Association of polygenic risk score (PRS) for schizophrenia and bipolar disorder with MMSE and education attainment.

| PRS | MMSE baseline<br>(n=20,276) |  | MMSE<24 at baseline<br>(n=20,276) |  | MMSE change<br>(n=4,248) |  | Ordinal Education<br>(n=79,480) |  |
| --- | --- | --- | --- | --- | --- | --- | --- | --- |
|  | Beta (95% CI)<sup>a</sup> | *P* | aOR (95% CI)<sup>a</sup> | *P* | Beta (95% CI)<sup>a</sup> | *P* | aOR (95% CI)<sup>b</sup> | *P* |
| Model 1: independent effect |  |  |  |  |  |  |  |  |
| SCZ | -0.063 (-0.095, -0.031) | <0.001 | 1.039 (1.037-1.040) | <0.001 | -0.082 (-0.160, -0.005) | 0.038 | 1.026 (1.012-1.041) | <0.001 |
| BPD | 0.040 (0.008, 0.072) | 0.013 | 0.945 (0.944-0.946) | <0.001 | 0.043 (-0.033, 0.119) | 0.272 | 1.014 (1.000-1.028) | 0.050 |
| Model 2: combined effect |  |  |  |  |  |  |  |  |
| Mean (SCZ, BPD) | -0.023 (-0.060, 0.015) | 0.232 | 0.977 (0.909-1.050) | 0.528 | -0.040 (-0.129, 0.049) | 0.381 | 1.040 (1.024-1.058) | <0.001 |
| Difference (SCZ - BPD) | -0.052 (-0.078, -0.026) | <0.001 | 1.058 (1.007-1.113) | 0.027 | -0.062 (-0.125, 0.000) | 0.050 | 1.006 (0.995-1.017) | 0.310 |
| Model 3: SCZ & BPD shared |  |  |  |  |  |  |  |  |
| SCZBPD vs. CONT | -0.010 (-0.040, 0.020) | 0.513 | 0.979 (0.924-1.036) | 0.459 | -0.094 (-0.166, -0.021) | 0.011 | 1.033 (1.020-1.047) | <0.001 |
| Model 4: SCZ-specific |  |  |  |  |  |  |  |  |
| SCZvs.BPD | -0.023 (-0.053, 0.007) | 0.131 | 1.010 (0.953-1.069) | 0.745 | -0.129 (-0.200, -0.057) | <0.001 | 0.984 (0.971-0.997) | 0.019 |

<sup>a</sup> Adjustment for gender, age, birth cohort, education level, TWB (chip version) and first 20 population stratification dimensions.
<sup>b</sup> Adjustment for gender, age, birth cohort, TWB (chip version), and first 20 population stratification dimensions.

When analyzing the MMSE as a binary outcome, the presence or absence of cognitive deficits, the results remained semilar. PRS_SCZ_ was associated with a higher risk of cognitive deficits (odds ratio [OR] per SD increase in PRS = 1.04, p < 0.001), while PRS_BPD_ was associated with a lower risk of cognitive deficits (OR = 0.95, p < 0.001). The difference between the PRS_SCZ_ and PRS_BPD_ was associated with a higher risk of cognitive deficits (OR = 1.06, p = 0.03).

For MMSE changes during follow-up, PRS_SCZ_ (beta = -0.08, p = 0.04) and the difference between PRS_SCZ_ and PRS_BPD_ (beta = -0.06, p = 0.05) were associated with decreased MMSE scores. PRS_BPD_ was not associated with changes in the MMSE scores. PRS_SCZ+BPD_ (beta = -0.09, p = 0.01) and PRS_SCZvsBPD_ (beta = -0.13, p < 0.001) predicted decreases in the MMSE scores.

For ordinal education, both PRS_SCZ_ (OR = 1.03, p < 0.001) and PRS_BPD_ (OR = 1.01, p = 0.05) were associated with higher educational attainment, but the difference (PRS_SCZ_ - PRS_BPD_) was not. Shared risk between SCZ and BPD (PRS_SCZ+BPD_) was positively associated with educational attainment (OR = 1.03, p < 0.0001), whereas SCZ-specific risk (PRS_SCZvsBPD_) was negatively associated with educational attainment (OR = 0.98, p = 0.02).

Sensitivity analyses for PRS thresholds of 0.5, 0.1, and 0.05 are shown in Supplementary Table 2-4. The pattern of results was similar.

Thin-plate splines for the estimated MMSE are shown in Figure 1. Compared with individuals with a low PRS_BPD_, the risk effect of PRS_SCZ_ on MMSE was larger in those with a high PRS_BPD_. Compared to individuals with high PRS_SCZ_, the protective effect of PRS_BPD_ on MMSE was larger in those with low PRS_SCZ_. There was a synergistic effect between PRS_SCZ_ and PRS_BPD_ on the MMSE scores. Both PRS_SCZ_concordant_ and PRS_SCZ_discordant_ scores were negatively associated with MMSE scores, and there was no synergistic effect between them. PRS_BPD_concordant_ was moderately negatively associated with MMSE scores in those with low PRS_BPD_discordant_, but PRS_BPD_concordant_ was moderately positively associated with MMSE scores in those with high PRS_BPD_discordant_. PRS_BPD_discordant_ was not associated with MMSE in those with low PRS_BPD_concordant_ but was positively associated with MMSE in those with high PRS_BPD_concordant_.

**Figure 1.**
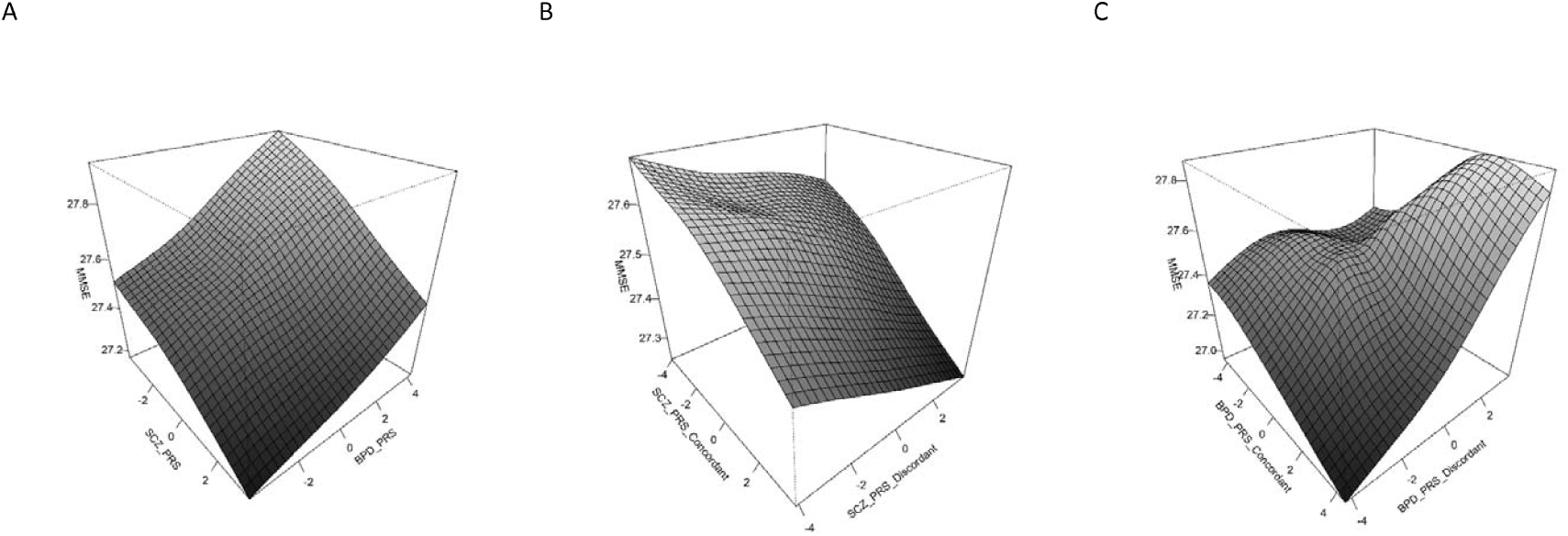
The thin-plate splines for estimated MMSE by (A) PRS_SCZ_ and PRS_BPD_ jointly (B) PRS_SCZ_concordant_ and PRS_SCZ_discordant_ jointly (C) PRS_BPD_concordant_ and PRS_BPD_discordant_ jointly.

## Discussion

Using a large collection of community samples from the Taiwan Biobank, this study explored the independent and joint effects of polygenic risks of SCZ and BPD on cognitive aging and educational attainment using individual genotype and phenotype data. We found that polygenic liability for SCZ was associated with an increased risk of cognitive aging. However, polygenic liability for BPD had a reduced risk. An analysis using thin-plate splines also confirmed these effects. The finding that a larger difference between PRS_SCZ_ and PRS_BPD_ was associated with a lower MMSE score further confirms the opposite effect of polygenic liability between SCZ and BPD. In the split model analyses, we found that both PRS_SCZ_concordant_ and PRS_SCZ_discordant_ were negatively associated with MMSE scores without a synergistic effect. PRS_BPD_disconcordant_ was moderately positively associated with MMSE scores in individuals with high PRS_BPD_cordant_ scores, and vice versa. In terms of MMSE change, the findings were largely consistent with those for MMSE baseline, except that the shared risk between SCZ and BPD and the SCZ-specific risk were both associated with MMSE decline. Both polygenic liabilities for SCZ and BPD were positively associated with high educational attainment. Analysis of the combined effect and shared polygenic risk of these two disorders further confirmed the positive associations. Of note, SCZ-specific polygenic risk was negatively associated with education. The positive association between polygenic liability and educational attainment may be determined by the shared part of BPD.

The association between the polygenic liability of SCZ and cognitive aging was consistent with previous epidemiological studies, which showed that patients with SCZ have an increased risk for developing dementia.^42^ Furthermore, one study found that first-degree relatives of probands with frontotemporal dementia have an increased risk of developing SCZ,^43^ indicating a shared genetic risk between SCZ and cognitive decline. In addition, several studies have demonstrated that the polygenic risk of SCZ is associated with poor cognitive performance, especially among older adults.^44-46^

Although previous epidemiological studies have also demonstrated that patients with BPD have a higher risk of developing dementia,^47^ we found that the polygenic liability of BPD was related to a high MMSE score at baseline. Previous studies showed inconsistent findings on the relationship between BPD and cognition. While some studies demonstrated polygenic liability for BPD might be associated with poor cognitive performance,^48^ another study showed that patients with BPD have higher cognitive abilities compared to controls.^49^ Of note, there were complex interactions between PRS_BPD_concordant_ and PRS_BPD_discordant_. PRS_BPD_disconcordant_ was moderately positively associated with MMSE scores in those with high PRS_BPD_cordant_ scores, and vice versa. However, PRS_BPD_concordant_ was modestly negatively associated with MMSE scores in those with low PRS_BPD_discordant_. There was no association between PRS_BPD_discordant_ and MMSE scores in those with low PRS_BPD_concordant_. These findings indicate that genetic factors have diverse effects on cognition. Further investigations are needed to determine the complex associations.

Among our study sample, 4280 patients had repeated MMSE measurements. The findings for the polygenic liability of SCZ were consistent with the analysis for MMSE baseline. However, the association of PRS_BPD_ was not significant, which might be due to the limited sample size. Notably, we found that SCZ-specific liability was associated with MMSE decline, in line with a recent study using GWAS summary data showing that SCZ-specific genetic background differentiated from BPD has a negative genetic correlation with general cognitive function.^21^ Taken together, these findings support the hypothesis that current SCZ diagnoses may aggregate into two subtypes: patients with high intelligence and BPD and those who show cognitive impairments independent of BPD.^22^

The finding that polygenic liability for both SCZ and BPD was positively associated with high educational attainment is consistent with previous studies demonstrating that there are overlapping genes between education and SCZ^50^ or BPD.^35^ However, the positive association of polygenic liability for SCZ with education might be attributed to the shared part with BPD. We found that the SCZ-specific risk, in contrast to BPD, was negatively associated with education. Our findings are consistent with the results of a recent study using linkage disequilibrium score regression showing that both SCZ and BPD genetic backgrounds have a positive but SCZ-specific genetic background that has a negative genetic correlation with educational attainment.^21^

Education and cognitive performance have a shared genetic basis.^51^ In addition, higher educational attainment could allow individuals to preserve better cognitive performance in late life.^52^ However, the association of genetic liability for SCZ and BPD for education attainment and cognitive aging was not consistent in our study. Cognitive aging might be determined by non-cognition factors, such as underlying cardiovascular diseases,^53^ which are commonly noted among individuals with SCZ or BPD.^54^

The PRS only considers common variants and thus does not capture the total heritability from family studies. For genetic architecture may differ across populations, using cross-ancestry GWAS results to calculate PRS may lead to a low prediction.^55^ When using European ancestry as discovery samples, the prediction performance in target samples of Asian or African ancestry was 37–78 % lower compared with that in target samples of European ancestry.^56^ In addition to the issue of cross-ancestry PRS prediction, the sample size for the discovery sample is also crucial. We used the most well-powered GWAS from PGC as discovery samples to identify susceptibility variants. Surprisingly, for predicting BPD in the Taiwan Biobank, PRS_SCZ_ and PRS_SCZ+BPD_ led to a better prediction than PRS_BPD_. There are two possible explanations. First, this implied that SCZ and BPD share a genetic architecture; the genetic correlation between them has been shown to be high (0.7–0.8).^57^ Second, this may be due to the fact that the case number for BPD GWAS (41 917 cases) is smaller than for SCZ GWAS (67 390 cases) and for SCZ + BPD GWAS (53 555 cases). In addition, the majority of samples for BPD GWAS are Caucasian and ∼20 % of the samples for SCZ GWAS are of East Asian ancestry.

Compared to PRS_BPD_, PRS_BPD_concordant_ explained a higher variance in SCZ. For predicting BPD, PRS_BPD_concordant_ explained more variance than PRS_BPD_ and PRS_SCZ_concordant_ explained more variance than PRS_SCZ_. Our results showed that splitting the PRS improved the prediction and provided genetic evidence that SCZ and BPD were not genetically homogeneous.

This study has several limitations. First, the disease status of patients with SCZ and BPD was obtained by retrospective self-reporting, which may have led to recall bias and resulted in misclassification and underestimation of the prevalence of diseases. A recent study^58^ has evaluated the accuracy of the self-reported disease status with the ICD diagnosis in the Taiwan National Health Insurance Research Database. The data show that the tetrachoric correlations for SCZ and BPD were 0.95 and 0.72, respectively. Second, we used the MMSE to detect cognitive aging; however, the MMSE is affected by education level. Patients who are well-educated perform better in the MMSE. In our study, we found that PRSscz and PRS_BPD_ were positively associated with educational attainment. Thus, the adverse effects of PRSscz on cognitive aging might be underestimated. Third, the Taiwanese Biobank participants may have been biased towards being healthy. Individuals with high genetic liabilities for psychiatric disorders and poor mental health had a low chance of being included in our analyses. Fourth, we only used individuals of Asian ancestry. Further investigation is warranted to determine whether our findings can be generalized to other populations.

This study provides evidence for the contrasting effects of polygenic liabilities in SCZ and BPD on cognitive aging. Although polygenic liabilities for both SCZ and BPD were independently associated with higher educational attainment, SCZ-specific polygenic liability was associated with lower educational attainment. Our findings partially support the hypothesis that the heterogeneity of SCZ and the positive association of polygenic liability for SCZ with education might be attributed to the shared part with BPD.

## Data Availability

GWAS summary results for BPD and SCZ are available online at

https://www.med.unc.edu/pgc

## Acknowledgements

This study was supported by the National Health Research Institutes (NHRI-EX109-10931PI, NHRI-EX110-10931PI, and NHRI-EX111-10931PI).

